# WHO TARGETS FOR CERVICAL CANCER CONTROL BY 2030: A BASELINE ASSESSMENT IN SIX AFRICAN COUNTRIES

**DOI:** 10.1101/2022.05.02.22274589

**Authors:** D Cristina Stefan, Jean-Marie Dangou, Prebo Barango, Issimouha Dille Mahamadou, Sharon Kapambwe

**Affiliations:** Institute of Global Health Equity Research Rwanda, University of Global Health Equity Rwanda; World Health Organization Regional Office for Africa, PO. Box: 06, Brazzaville, Congo Republic

## Abstract

**Aim:** We present and analyze the findings of a survey of the readiness of the healthcare systems in Eswatini, Guinea, Malawi, Rwanda, Uganda and Zambia, to implement the necessary measures for attaining the targets for cervical cancer control, set by WHO, by the year 2030.

**Methods:** A questionnaire with 129 questions with preset answer options was completed in 2020, by ministries of health program coordinators for non-communicable diseases, cancer control and/or reproductive health, and by WHO country offices, in the six countries selected.

**Results:** The findings on demographics, burden of disease, governance and management, laboratory services, equipment, supplies and medicines, as well as on personnel and training will be presented here.

The burden of cervical cancer in the countries studied is considerable, based on the IARC estimations. The incidence of the disease is augmented by the high prevalence of HIV infection, in most of the countries surveyed. Most of the population live in rural areas, where access to the health services is far from ideal. Facilities for screening with HPV tests and for histopathology are limited. One pathologist covers the diagnostic needs of between 0.5 million and 4 million inhabitants. Most other categories of health professionals are under-represented, and the capacity to train them is inadequate.

**Conclusions:** Strong country commitment and leadership, innovative solutions and extensive international cooperation would be needed to attain the targets of cervical cancer control set by WHO, in these countries.

## Introduction

The International Agency for Research in Cancer (IARC) estimated that in 2020, close to 342,000 women died from cancer of the uterine cervix, in the world. The age-standardized mortality from the disease was 17.4/100,000 in low-income populations, seven times higher than the 2.5/100,000 recorded in the high-income populations. In the same year, according to IARC, around 603.300 new cases of cervical cancer appeared woldwide^1^.

According to IARC predictions, the above figure will grow to 800,000 by 2040. A large share of these new cases will appear in low- and low mid-income countries. The African continent would see its number of incident cases almost double in the next 20 years, from 117,000 to 228,000^2^.

By implementing adequate measures for cervical cancer control, this ascending trend can be reversed. It is already possible to prevent cervical cancer by vaccinating young girls against HPV, before the onset of sexual activity^3^, and by screening women for precancerous changes, to remove those, if diagnosed. Cervical cancers detected and treated in early invasive stages have high rates of cure^4^. The difficulties on the way to implementing the measures mentioned are substantial but can be overcome. The reward would be that sometime in the future, the cancer of the cervix will be relegated to the books of history of medicine.

In 2018, the Director-General of WHO made a call to action for elimination of cervical cancer as a public health problem. To achieve this aim, the world-wide incidence of the disease should be brought below 4/100,000. A global strategy towards that goal was adopted by the World Health Assembly in 2020. A first target, to be attained by 2030, consists of: world-wide vaccination of 90% of girls by 15 years of age, screening of 70% of women with a high performance test for detection of pre-cancerous lesions of the cervix, by the age of 35, and again by the age of 45, treating 90% of the detected pre-invasive cancers and managing 90% of women with invasive cancers^5^.

The NCD division of WHO/AFRO selected to support a “first wave” of six countries, in their effort to achieve the above targets: Eswatini, Guinea, Malawi, Rwanda, Uganda and Zambia. The experience gained in the process will be used to extend the support to other countries on the continent.

We report the findings of an evaluation by AFRO, of the readiness to implement and to scale up the measures necessary for the achievement of the targets set for 2030, in the first wave countries. This is the first part of the report, where only the findings concerning demographics, burden of disease, governance and management, laboratory services, equipment, supplies and medicines, as well as personnel and training will be presented. The remaining of the data, dealing with the state of primary and secondary prevention, of the treatment with curative intent and the state of palliation, will be published soon.

## Methods

A detailed questionnaire in English was mailed in May 2020 to national program coordinators for non-communicable diseases, cancer control and/or reproductive health national programs coordinators, ministries of health, representatives and WHO country offices. The officials collaborated with each other in drawing the answers, resulting in a single completed questionnaire for each country.

The 129 questions provided preset answer options. Further consultations were held via internet for omitted questions or answers requiring clarification. Consultations were held in English, except for Guinea representative which were held in French. The results were compiled in comparative tables as shown below

## Results

### 1. Demographics and Epidemiology

**Table I:**
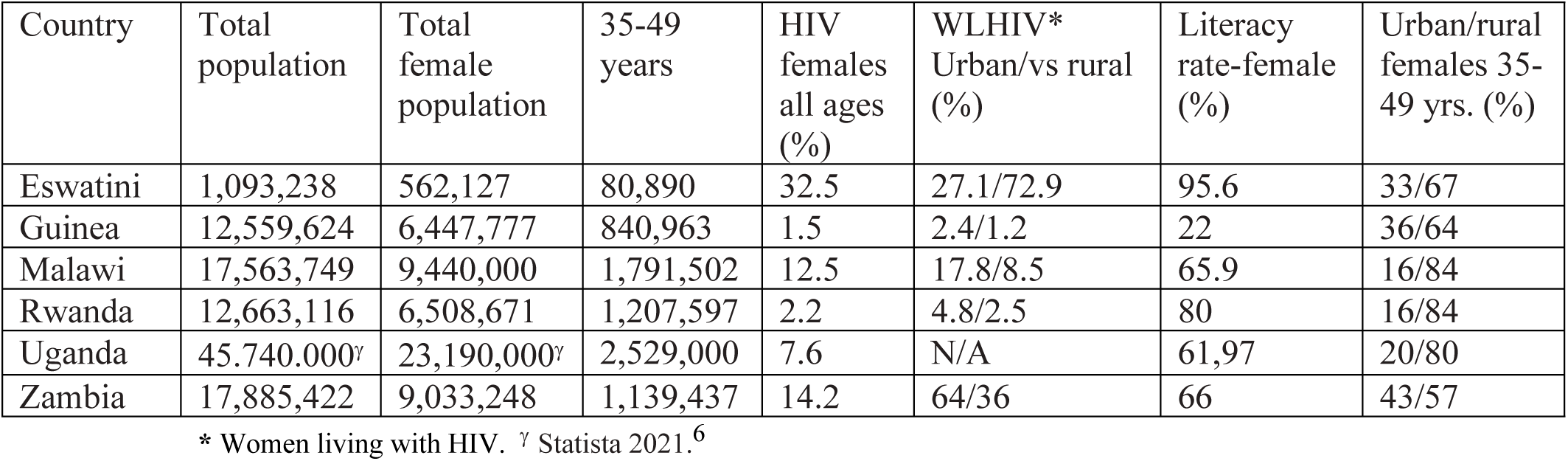
Demographics

| Country | Total population | Total female population | 35-49 years | HIV females all ages (%) | WLHIV* Urban/vs rural (%) | Literacy rate-female (%) | Urban/rural females 35-49 yrs. (%) |
| --- | --- | --- | --- | --- | --- | --- | --- |
| Eswatini | 1,093,238 | 562,127 | 80,890 | 32.5 | 27.1/72.9 | 95.6 | 33/67 |
| Guinea | 12,559,624 | 6,447,777 | 840,963 | 1.5 | 2.4/1.2 | 22 | 36/64 |
| Malawi | 17,563,749 | 9,440,000 | 1,791,502 | 12.5 | 17.8/8.5 | 65.9 | 16/84 |
| Rwanda | 12,663,116 | 6,508,671 | 1,207,597 | 2.2 | 4.8/2.5 | 80 | 16/84 |
| Uganda | 45,740,000<sup>γ</sup> | 23,190,000<sup>γ</sup> | 2,529,000 | 7.6 | N/A | 61.97 | 20/80 |
| Zambia | 17,885,422 | 9,033,248 | 1,139,437 | 14.2 | 64/36 | 66 | 43/57 |

\* Women living with HIV. <sup>γ</sup> Statista 2021.<sup>6</sup>

Most women live in rural areas in all six countries. The numbers of HIV infected females vary within a broad range, from 1.5% in Guinea to 32.5% in Eswatini. The female literacy rate varied from 22 % in Guinea to 95.6% in Eswatini.

**Table II:** Burden of disease and registries available

| CANCER TODAY (ASR - World) in 2020 |  |  | Country registries |  |  |  |
| --- | --- | --- | --- | --- | --- | --- |
| Country | Incidence invasive CxCa (/100,000 per year) | Mortality (/100,000 per year) | Cancer registry | Registry of invasive cervical cancer | Registry of screening and precancerous lesions | Registry of immunized for HPV |
| Eswatini | 84.5 | 55.7 | YES | NO | At the units | NO |
| Guinea | 50.1 | 37.2 | YES | NO | At hospital | NO |
| Malawi | 67.9 | 51.5 | YES | NO | At national level | YES |
| Rwanda | 28.2 | 20.1 | YES | NO | At the units | NO |
| Uganda | 13.3 | 41.4 | YES (Kyadondo district) | NO | YES | YES |
| Zambia | 66.4* | 44.5* | YES (Lusaka district) | At units | At units | NO |

\* Data from 2018<sup>7</sup>

Except Uganda and Rwanda, the age-standardized incidence of cervical cancer in the countries surveyed is two to three times higher than the average figure for Africa, which is 25.6 /100,000. The mortality figures, excluding Rwanda, are also twice or even three times higher than the average of 17.7/100,000 for the whole continent^8^.

### 2. Governance and management

**Table III.** Governance and management. Cervical cancer prevention and control programs.

| Country | Health policy, plan, strategy | MOH cervical cancer existing unit | NCCP | National policy, plan or strategy eHealth | Unit e-Health | HPV vaccination program/unit | Cervical cancer policy, plan or strategy | Guide-lines | National M&E plan CCPN | MOH section, unit or team M&E |
| --- | --- | --- | --- | --- | --- | --- | --- | --- | --- | --- |
| Eswatini | YES | YES | YES | NO | NO | YES | YES | YES | YES | NO |
| Guinea | NO | YES | NO | YES | YES | NO | NO | YES | YES | NO |
| Malawi | YES | YES | YES | YES | YES | YES | YES | YES | YES | N/A |
| Rwanda | YES | YES | YES | YES | YES | YES | YES | YES | YES | YES |
| Uganda | YES | YES | YES | YES | YES | YES | NO | NO | NO | NO |
| Zambia | YES | YES | YES | N/A* | N/A | YES | YES | YES | N/A | N/A |

\*N/A: data not available

Most of the first wave countries have national cancer control plans, as well as cervical cancer control plans and guidelines, drawn according to WHO guidelines. However, although provisions for monitoring and evaluation of the activities planned are made, a specific team tasked with monitoring and evaluation exists only in the Rwandan Department of Health.

### 3. Laboratories

**Table IV.** Laboratories and pathologists

| Country | National policy plan or strategy lab. | Lab. accreditation Plan | Lab. system centralized | National reference lab. | PUBLIC |  | PRIVATE |  |
| --- | --- | --- | --- | --- | --- | --- | --- | --- |
|  |  |  |  |  | Labs | Pathologists | Labs | Pathologists |
| Eswatini | NO | NO | YES | YES | 1 | 1 | 2 | 1 |
| Guinea | NO | NO | NO | YES | 2 | 3 | 0 | 0 |
| Malawi | YES | YES | YES | YES | 4 | 4 (incl. private) | 2 | 4 (incl. public) |
| Rwanda | YES | YES | YES | YES | 5 | 15 (incl. private) | 1 | 15 (incl. public) |
| Uganda | YES | YES | YES | YES | 4 | 5 | 10 | 10 |
| Zambia | YES | NO | YES* | YES | 5 | 15 (incl. private) | 5 | 15 (incl. public) |

\* At regional level

The above data, correlated with the respective countries’ population, indicate a ratio of one pathologist for between 0.5 and 4 million inhabitants, depending on country.

Laboratories will need to ensure the transition of screening for cervical cancer from visualization after application of acetic acid (VIA) or Papanicolaou smear to testing for Human Papillomavirus (HPV). The current state of HPV testing and histopathology is shown below.

**Table V.** State of HPV testing and histology

| Country | Providing HPV testing |  |  | HPV results recorded | Challenges to cervical cancer histology |
| --- | --- | --- | --- | --- | --- |
|  | Public | Private | NGO |  |  |
| Eswatini | 0 | 0 | 0 | 0 | N/A |
| Guinea | 0 | 0 | 0 | 0 | N/A |
| Malawi | 0 | 1 | 2 | N/A | Under research (data not available) |
| Rwanda | 14 | 0 | 0 | NO | Lacking machinery and consumables |
| Uganda | 11 | 2 | 1 | NO | N/A |
| Zambia | 0 | 1 | 1 | NO | Old histopathology equipment frequently breaking down; poor supply of consumables |

The present state of the pathology laboratories, combined with the paucity of pathologists, requires urgent action.

### 4. Equipment, supplies and medicines

Cryotherapy, thermal ablation and LLETZ equipment is available in most countries surveyed; there is no LLETZ in Uganda and no thermal ablation in Eswatini. Neither cervical cancer chemotherapy drugs nor opiates are available in Guinea.

**Table VI.**
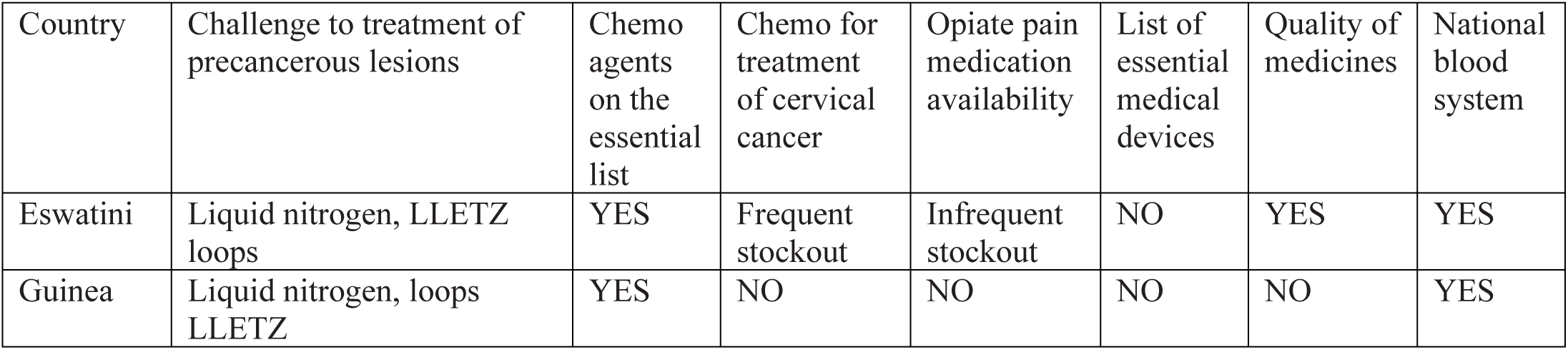
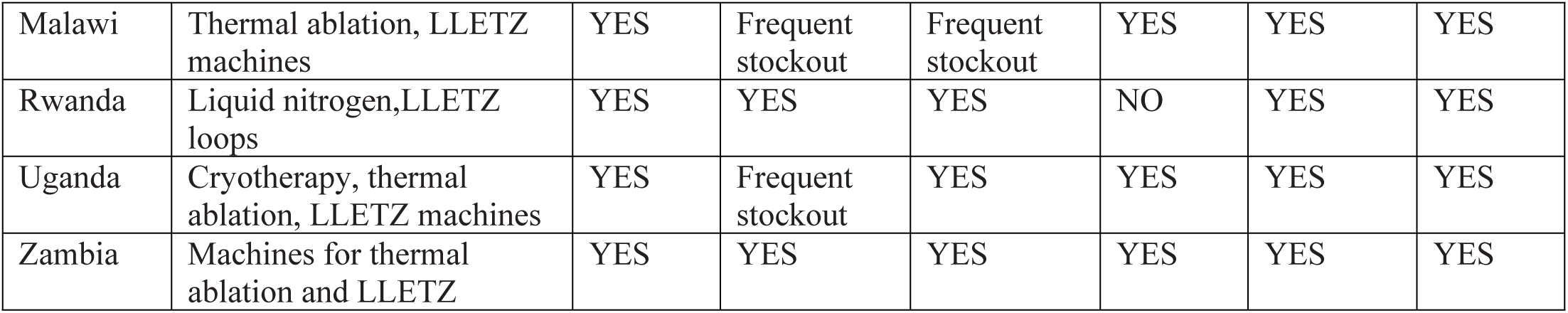
and Challenges to the treatment of precancerous lesions; equipment and medicines availability.

**Table VII.** Radio diagnostic and radiotherapy machines

| Country | Radiodiagnostic |  |  | Radiotherapy |  |  |
| --- | --- | --- | --- | --- | --- | --- |
|  | CT<br>(total/functional) | MRI<br>(total/functional) | PET | Linear<br>accelerator | Telecobalt | Brachytherapy |
| Eswatini | 2/? | 1 | 0 | 0 | 0 | 0 |
| Guinea | 5/3 | 2/1 | 0 | 0 | 0 | 0 |
| Malawi | 3/3 | 1/0 | 0 | 0 | 0 | 0 |
| Rwanda | 10/9 | 2/2 | 0 | 2/2 | 0 | 0 |
| Uganda | 10/7 | 3/1 | 0 | 1 | 2 | 1 |
| Zambia | 16 | 3 | 0 | 1 | 2 | 2 |

An absence of PET scans and a paucity of brachytherapy facilities require action soon. Moreover, both maintenance and repair of the existing equipment need to be improved.

### 5. Personnel and training

**Table VIII.**
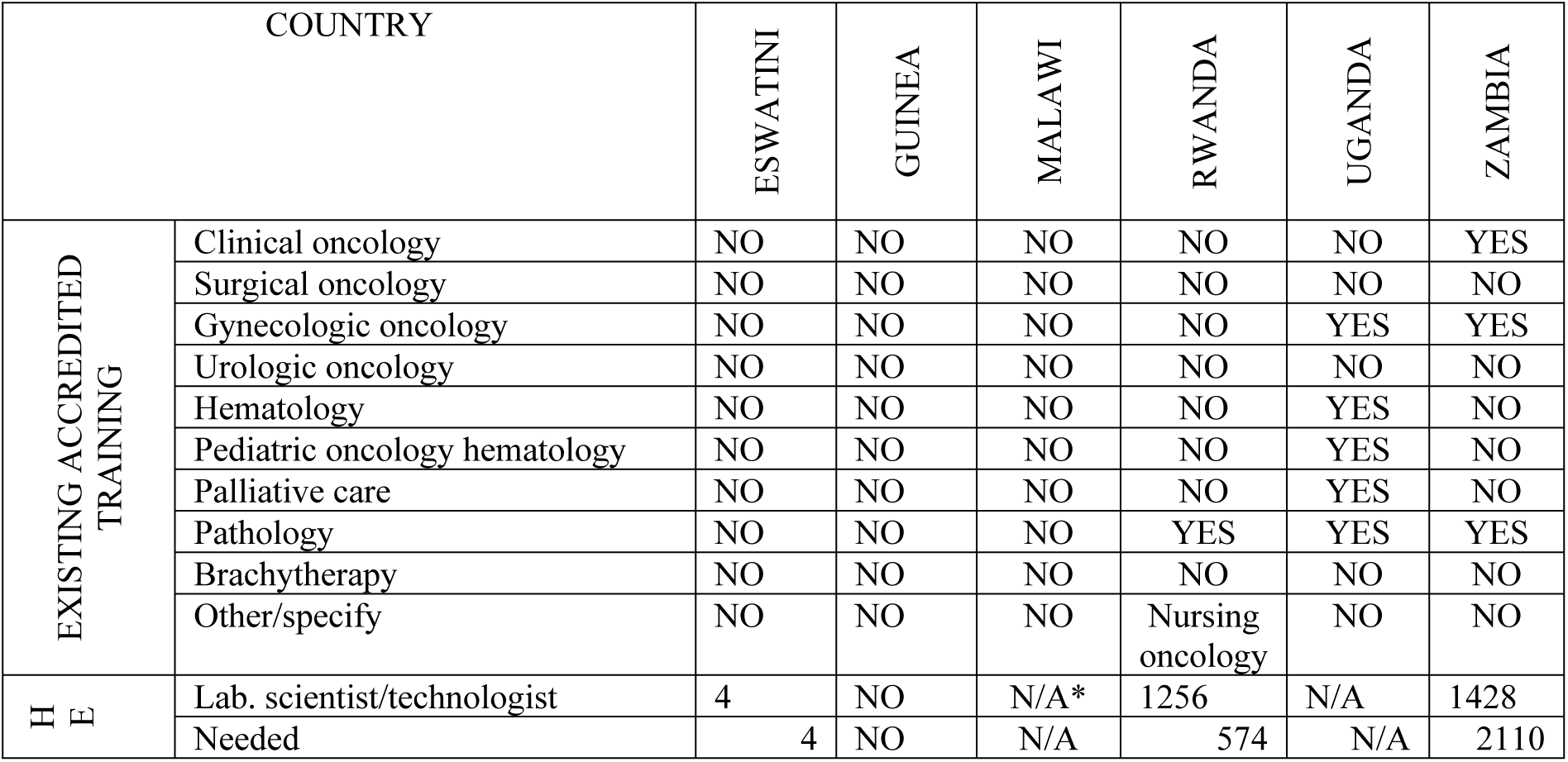
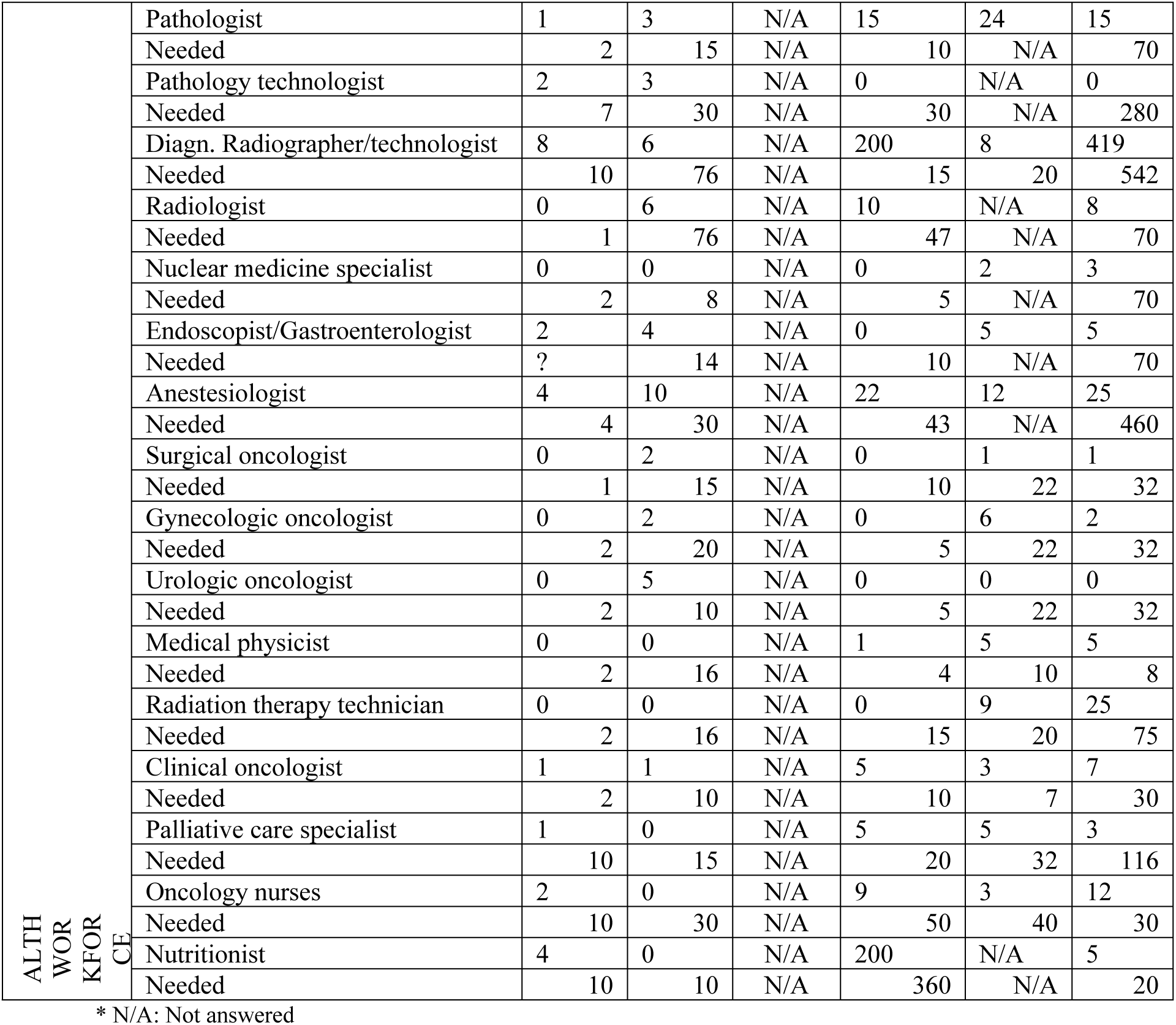
Training possibilities and estimated need of professionals in the field of cancer care.

There is a great need for training of super-specialists in oncology fields, which at this time is not possible in most of the first wave countries. Only Uganda and Zambia have programs for training gynecological oncologists. Pathologists, crucial for the diagnosis and management plan of cancers, are only being trained in Rwanda, Uganda and Zambia.

Zambia estimates the highest need of professionals in almost all fields related to oncology. Malawi did not provide an answer to this question. Uganda did not report a definite need for some of the professions.

## Discussion

### Cancer registration

The first wave countries cumulatively have a female population of almost 55,200,000. In 2020, it is estimated that 17,903 women were diagnosed with cervical cancer there, and 11,922 women died. It is important to be aware that these reported estimations are provided by IARC and not by country cervical cancer registries. Data on invasive cervical cancer, screening for pre-invasive lesions or on vaccination are not centralized. For realistic planning by the Health Ministries, including an efficient allocation of resources, reliable national cervical cancer data registration should be available.

### Governance and management

Most first wave countries report having national cancer control plans, except for Guinea. Apart from Uganda and Guinea, all countries have cervical cancer control plans and guidelines. However, there is a clear need to institute a mechanism for evaluation of the results of those plans and guidelines in practice. Of the six respondents, only Rwanda had a cervical cancer control monitoring team at governmental level.

It is remarkable that four of the countries surveyed have governmental measures in place to develop e-Health facilities; they have e-Health units as well. The role of medical telecommunication may prove a determinant factor for the success of fighting cervical cancer: it will make it easier to centralize and assess data on the disease burden, to organize doctor-to-doctor consultations on difficult cases, as well as online tumor boards, to access scientific information for clinical decision support and to consult guidelines. As it will be shown in the next section, the telepathology facility will be indispensable for the diagnosis and management of cervical cancer in Africa.

As part of the e-Health activities, the m-Health (via mobile telephone) could be leveraged to deliver information on the disease, as well as on the HPV vaccination and screening, and maintain in the system the patients, by means of appointments, reminders or navigation of the referral pathways, for the whole duration of their management.

### Laboratories and pathologists

The ratios of laboratories and pathologists to population in the first wave countries are not reassuring. The figures for pathologists are far behind the best ratios in Africa, of between 1/50.000 and 1/200.000^9^, registered in Algeria and Tunisia. In UK, where the density of pathologists is 1 to little under 30,000 inhabitants, a survey published in 2018 by The Royal College of Pathologists^10^ found that 78% of the departments surveyed needed more consultants. Moreover, pathologists do not work alone; their task is facilitated by medical technologists or technicians, histo- and cyto-technologists, information technology specialists, forensic pathology technicians, administrative and clerical personnel, and others. It can be easily inferred that the pathology workforce in the countries surveyed by us would require a considerable reinforcement.

The cervical cancer screening targets set for WHO for 2030 require large scale shifting from inspection of the cervix with acetic acid or Pap smear to HPV testing. This testing is automatized; however, besides acquiring the machines capable of doing it, the suitable infrastructure, including a reliable provision of clean water, electricity and distance communication, as well as personnel trained to prepare the samples for analysis, service the machine and attend to possible breakdowns, should be available.

Additionally, at present, HPV testing methods are not well suited for a test-and-treat approach, hence the need for an efficient follow-up system for women identified as high-risk.

Further, the scaling up of screening would generate considerably more cytology smears and cervical biopsies, hence a substantial increase in demand for cytopathological and histopathological diagnoses. With the 18 full pathology courses offered in Africa at the present, it was calculated that only in several centuries will Sub-Saharan Africa attain the present ratios of pathologists to population found in US or UK^9^. Besides training personnel abroad, the acquisition of slide scanners, connected to the Internet, should be imperiously considered. That would make possible the outsourcing of slide reading, either to human pathologists or to artificial intelligence systems (AI). For image evaluation, AI is as performant as humans or even better, to the point where in a not too far future it may be considered unethical not to use it when examining a slide^11^.

### Equipment, supplies and medicines

Guinea reported a complete absence of cervical cancer chemotherapy drugs and opiates. An in-depth analysis of the causes could not be conducted at the time of the survey.

Another significant finding is that brachytherapy is available only in Zambia and Uganda. Eswatini and Malawi reported no radiotherapy facilities for cervical cancer. Given that most patients contact the health institutions late in the course of the disease, when surgery is not indicated, and radiation or chemo-radiation are the indicated therapies, many women might not have access to proper treatment in these countries. To attain the 90% rate of treatment envisaged by WHO by 2030, it would be necessary to build more radiotherapy facilities, including brachytherapy. The initial costs for a radiation bunker and brachytherapy machine, including the radioactive sources for the first year would amount to around 1.3 million US dollars^12^, while a new linear accelerator may cost up to 2.8 million dollars.

### Personnel and training

The data in Table VII show a general shortage of personnel, in all professions related to the prevention and treatment of cervical cancer, in the countries studied. It is perhaps useful to highlight the overall need for 191 radiologists, 537 anesthesiologists, 663 radiographers, 2688 laboratory technologists, as more salient figures. And these figures are only for the six countries surveyed.

The methodology used to assess these needs is, however, unknown. The complexity of planning the healthcare human resources needed in the future was described, without giving definitive solutions, repeatedly in the literature ^13, 14, 15^. However daunting that task may be, it needs to be accomplished in the countries studied, as a first step towards attaining the targets set by WHO for 2030.

Training the doctors, nurses and other medical personnel mentioned in Table VIII will be probably not possible inside the countries studied, of which some do not have any training facilities. Some of the people aspiring to enter these professions might be prepared to undergo their training abroad: the existing agreements with various academic institutions in other countries or continents, need to be revised and updated, while new agreements may need to be entered, to cover as much as possible the needs. This activity should take place within the frame of a ministerial human resources for health strategic plan.

At the same time, the governments of the countries surveyed may realize, based on a thorough analysis of personnel needs, that the opening or upgrading of national institutions of training in health professions is long overdue and needs to happen. However, given the long duration of training of specialists in gynecological oncological surgery, radiotherapy, pathology, anesthesia and intensive care – to mention just a few - interim solutions, aimed at the 2030 targets, might need to be be found.

As emphasized above, in this context, automatization of HPV testing and tele-histopathology appear to be a necessity, and not a luxury. Further, agreements could be concluded with medical schools from other African countries or even from overseas countries, to organize supervised training stages for their Master students, in the countries surveyed by us. All the same, young specialists from foreign countries, particularly from Europe, may be recruited and accredited to work for limited periods, in areas of need. The benefit would be mutual: young specialists would be exposed to developing countries’ health and healthcare characteristics and gain valuable experience, while the population would benefit from improved prevention and treatment of cervical cancer.

### What is new in this report?

This is an analysis of a survey by e-mailed questionnaire, with follow-up telephone interviews where necessary, aimed at evaluating the readiness of six African countries to achieve the targets formulated by WHO regarding the reduction of the burden of cervical cancer by 2030. The respondents were high authorities from national health administrations and WHO country representatives, thus ensuring the veracity of the data published here.

### What are the shortcomings of this study?

The data collated here are only as good as the primary sources were. The most obvious example is that in the absence of population based cancer registries and well-coordinated national cancer (or cervical cancer) registries, the figures for incidence and mortality reported were those estimated by IARC. Another shortcoming was that some questions were not answered.

## Conclusions

The burden of cervical cancer in the countries studied is considerable, based on the IARC estimations. The incidence of the disease is augmented by the high prevalence of HIV infection, in most of the countries surveyed. Most of the population lives in rural areas, where access to the health services is far from ideal. Facilities for screening with HPV DNA tests are severely limited, and histopathology services are insufficient. One pathologist is supposed to cover the diagnostic needs of between 0.5 million and 4 million inhabitants.

Most other categories of health professionals are under-represented or absent. Strong country commitment and leadership, innovative solutions and extensive sub-regional and international cooperation would be rather urgently needed to attain the targets of cervical cancer control set by WHO, in these countries.

## Data Availability

N/A

## REFERENCES

1. IARC Cancer Today. Cervix uteri. Available from: https://gco.iarc.fr/today/data/factsheets/cancers/23-Cervix-uteri-fact-sheet.pdf Accessed on 3 Jan. 2022

2. IARC. Cancer Tomorrow. Cervix uteri. Available from: https://gco.iarc.fr/tomorrow/en/dataviz/isotype?cancers=23&single_unit=5000&populations=324_454_646_748_800_894&group_populations=1&multiple_populations=1&sexes=2&years=2040 Accessed on 3 Jan. 2022

3. Torjesen I. HPV vaccine cut cervical cancer rates in England by 87%. BMJ. 2021 Nov 5;375:n2689. https://doi.org/10.1136/bmj.n2689

4. Wright JD, Matsuo K, Huang Y, Tergas AI, Hou JY, Khoury-Collado F, et al. Prognostic Performance of the 2018 International Federation of Gynecology and Obstetrics Cervical Cancer Staging Guidelines. Obstet Gynecol. 2019 Jul;134(1):49–57.

5. WHO. Global strategy to accelerate the elimination of cervical cancer as a public health problem [Internet]. [cited 2022 Feb 1]. Available from: https://www.who.int/publications-detail-redirect/9789240014107

6. Uganda - total population by gender 2020 [Internet]. Statista. [cited 2022 Jan 11]. Available from: https://www.statista.com/statistics/967968/total-population-of-uganda-by-gender/

7. Costing the National strategic Plan on Prevention and Control of Cervical Cancer: Zambia, 2019-2023 Geneva: World Health Organization; 2020. Licence: CC BY-NC-SA 3.0 IGO.

8. IARC. Cancer Today. Cervix uteri. Available from: https://gco.iarc.fr/today/online-analysis-table?v=2020&mode=cancer&mode_population=continents&population=900&populations=903&key=asr&sex=2&cancer=39&type=0&statistic=5&prevalence=0&population_group=0&ages_group%5B%5D=0&ages_group%5B%5D=17&group_cancer=1&include_nmsc=1&include_nmsc_other=1#collapse-group-1-0-0. Accessed on 3 Jan. 2022

9. Wilson ML, Fleming KA, Kuti MA, Looi LM, Lago N, Ru K. Access to pathology and laboratory medicine services: a crucial gap. Lancet. 2018 May 12;391(10133):1927–38. 10.1016/S0140-6736(18)30458-6

10. The Royal College of Pathologists. Meeting Pathology Demand. Histopathology workforce census. Published by The Royal College of Pathologists, London, UK. 2018. Available from https://www.rcpath.org/uploads/assets/952a934d-2ec3-48c9-a8e6e00fcdca700f/Meeting-Pathology-Demand-Histopathology-Workforce-Census-2018.pdf Accessed 17 Dec. 2021

11. Machine learning will replace human radiologists, pathologists, maybe soon [Internet]. Healthcare IT News. 2017 [cited 2022 Jan 3]. Available from: https://www.healthcareitnews.com/news/machine-learning-will-replace-human-radiologists-pathologists-maybe-soon

12. Williams TR. Starting a Brachytherapy Program in the United States. [Internet] Available from: http://www.aoic.net/elekta/elk1402archive/E204Williams.pdf Accessed on 15 Dec. 2021.

13. Lopes MA, Almeida AS, Almada-Lobo B. Handling healthcare workforce planning with care: where do we stand? 2015, Human Resources for Health 13(38) 10.1186/s12960-015-0028-0

14. Health Workforce Advocacy Initiative (HWAI). Guiding Principles for National Health Workforce Strategies. 2008. Available from: https://www.who.int/healthsystems/round9_6.pdf Accessed on 28 Dec. 2021

15. Moat KA, Waddell K, Lavis JN. Evidence Brief: Planning for the Future Health Workforce of Ontario. 2016 McMaster Health Forum Available from: https://www.mcmasterforum.org/docs/default-source/product-documents/evidence-briefs/workforce-planning-eb.pdf?sfvrsn=cf5455d5_3 Accessed 19 Dec. 2021

